# Genetic Risk for Alcohol Use Disorder in Relation to Individual Symptom Criteria: Do Polygenic Indices Provide Unique Information for Understanding Severity and Heterogeneity?

**DOI:** 10.1101/2024.09.20.24313762

**Authors:** Yongguk Kim, Sean P. Lane, Alex P. Miller, Kirk C. Wilhelmsen, Ian R. Gizer

**Affiliations:** Department of Psychological Sciences, University of Missouri-Columbia; Department of Psychiatry, Indiana University School of Medicine; Rockefeller Neuroscience Institute, West Virginia University; Department of Neurology, West Virginia University

**Keywords:** polygenic risk score, alcohol use disorder, genetic heterogeneity, severity, individual symptoms

## Abstract

Alcohol Use Disorder (AUD) is a heterogenous category with many unique configurations of symptoms. Previous investigations of AUD heterogeneity using molecular genetics methods studied the association between genetic liability and individual AUD symptoms at the latent level or focusing on a small number of genetic variants. Notably, these studies did not investigate potential severity differences between symptoms in their genetic analyses. Therefore, the current study aimed to examine the genetic risk for individual AUD symptom criteria by using a polygenic risk score (PRS) approach to assess the relative severity of each AUD symptom and test for associates with AUD symptoms above and beyond a unidimensional AUD construct. An AUD PRS was created using summary statistics obtained from published genome-wide association studies (GWAS), and Multiple Indicators Multiple Causes (MIMIC) models were employed to examine the effect of the PRS on overall AUD severity as well as on individual symptoms after accounting for this overall effect. The phenotypic severity of AUD symptoms was highly correlated with the genetic severity of AUD symptoms (*r* = 0.78). Results of MIMIC models indicated that the AUD PRS significantly predicted the AUD factor. Regression paths testing the unique, direct effects of the PRS on individual AUD symptoms, independent of the latent AUD factor, were not significant. These results imply that PRSs derived from GWAS of AUD influence symptom expression through a single genetic factor that is highly correlated with the relative severity of individual symptoms when measured at the phenotypic level. Item-level GWAS of AUD symptoms are needed to further parse heterogeneous symptom expression and allow for more nuanced tests of these conclusions.

---

Alcohol Use Disorder (AUD) is a highly heritable disorder [1] resulting in a varying collection of negative consequences spanning health, social, and interpersonal problems [2,3]. Although the Diagnostic and Statistical Manual of Mental Disorders, 5^th^ edition (DSM-5), currently describes AUD as a uni-dimensional disorder [4], recent studies [5,6] have suggested that AUD might be more heterogeneous with distinct symptoms potentially resulting from separable, though related, underlying risk mechanisms. Further, some previous studies [7] have suggested that this heterogeneity can be observed at the genetic level. These studies indicate that meaningful information is lost when aggregating distinct AUD symptoms into a single, dichotomous category. If so, examining individual AUD symptoms is likely to give us valuable insight into the etiology of AUD. Thus, the current study aimed to examine the genetic risk associated with individual AUD symptom criteria.

Animal models of alcohol use clearly demonstrate distinct genetic liabilities underlying different alcohol use phenotypes, including individual AUD symptoms. For example, studies have shown that drosophila can be independently bred to show more or less alcohol consumption, more or less susceptibility to alcohol’s sedating effects [8], and higher or lower rates of relapse after periods of alcohol deprivation [9]. Selective breeding studies using rodents have yielded similar evidence in terms of alcohol drinking preference [10,11] and susceptibility to seizures due to alcohol withdrawal [12]. These studies have also suggested that there might be separable genetic influences responsible for individual AUD symptoms such as alcohol *tolerance* and *withdrawal* [13-15].

These findings have been supported by behavior genetics studies conducted in human samples. Early twin studies indicated that some AUD symptoms might be more genetically influenced (e.g., *withdrawal*) than others (e.g., *continued use*) [16,17]. More recently, a twin study showed that each symptom differs in terms of its heritability with a range from 36% (*desire to quit*) to 59% (*time spent*) [7], and a molecular genetic study demonstrated that single nucleotide polymorphism (SNP) heritability estimates of individual symptoms ranged from 13% to 39% [18,19]

Behavior genetic studies have also been used to explore the potentially multi-factorial structure of AUD symptomatology. For example, Kendler and colleagues [7] found evidence that the genetic liability for AUD might consist of three separable factors, which they labeled tolerance, loss of control with social dysfunction, and withdrawal. Furthermore, early molecular genetic studies using linkage analysis have noted heterogeneity in the genetic etiology of individual symptoms. For example, one study found that the relation between a chromosome 4 locus and AUD was specific to symptoms of *tolerance* and drinking larger amounts or for a longer period of time than intended (*larger/longer)* [20], and a second study reported evidence of linkage to a chromosome 9 region specific to the symptom of *withdrawal* [21]. More recent genome-wide association studies (GWAS) have found distinct genetic loci related to AUD symptoms such as *craving* [22], the inability to cut down or stop drinking (*cut/stop*) [23], *time spent* drinking, *larger/longer*, and *tolerance* [24]. Finally, studies of the alcohol dehydrogenase (ADH) genes central to the metabolism of alcohol have shown that variation in these genes predicted specific AUD symptoms in both European [25,26] and African ancestry samples [27].

Evidence of heterogeneity in the presentation of AUD and the etiology of its symptoms can be observed in phenotypic human studies as well. According to DSM-5, only two AUD symptoms are needed to meet the diagnostic criteria, which yields 2,037 theoretically possible combinations of symptoms, with 55 possible combinations for two symptoms alone. Indeed, endorsement rates differ across the individual symptoms, forming a smaller number, though hundreds of AUD profiles have been observed in epidemiologic studies [28]. These different profiles of AUD symptoms are likely to be associated with differences in alcohol consumption patterns, alcohol-related consequences, correlates of AUD, and progression to more severe forms of AUD [28–30]. Further, although variable-centered studies using factor analysis have shown that AUD symptoms can be represented by a single factor [e.g., 31], person-centered studies using cluster analysis or latent class analysis have suggested that there might be multiple sub-factors underlying AUD symptomatology [e.g., 32]. Consistent with findings from the behavior genetics study described above [7], one recent study [6] used factor analysis and hierarchical clustering analysis to evaluate the structure of AUD and found evidence supporting a multi-dimensional conceptualization of the disorder, indicating a three-factor solution: 1) tolerance and excess consumption, 2) withdrawal, and 3) loss of control. A similar study using latent class analysis found a similar three-factor solution [5]. Results from both studies are consistent with the structure described above at the genetic level [7], thus suggesting this multidimensional structure is observable at multiple levels of analysis.

Notably, different AUD profiles might not simply reflect a multi-factorial underlying structure, but could also reflect differences in AUD severity that go beyond differences in symptom count to predict meaningful differences in experienced negative consequences associated with AUD. At the level of individual symptoms, Item Response Theory (IRT) has been used to investigate the relations of AUD symptoms to a single underlying latent trait indexing AUD symptomatology. For example, research has shown that *tolerance* is a frequently endorsed symptom reflecting a lower level of AUD severity relative to other symptoms, whereas *withdrawal* is infrequently endorsed and reflects a higher level of severity [33,34]. Further, the study by Bailey and his colleagues [5] referenced above that used latent class analysis to identify AUD symptom clusters also reported that these symptom clusters were associated with differences in AUD severity (consumption < loss of control < withdrawal). Thus, these results suggest that some of the heterogeneity in symptom presentation could be a reflection of symptom clusters representing distinct features of AUD, but they also suggest that these features are ordered in terms of AUD severity. As these conclusions are not mutually exclusive and can be difficult to differentiate empirically, there continues to be a rich debate in the literature regarding the potential multidimensionality of AUD.

Taken together, the reviewed studies imply that item-level genetic investigations of AUD symptoms are warranted and have the potential to expand our understanding of AUD’s etiology. For example, studies have yet to investigate whether symptom criteria index AUD severity at the genetic level. Further, previous studies have shown that AUD symptoms may have distinct genetic etiologies, but these studies have largely been limited to examinations of their heritability [e.g., 7,19]. A smaller number of association studies have examined these differences at the level of measured genotypes but have either relied on a candidate gene approach or been limited by low statistical power by GWAS standards [e.g., 24, 27].

To address the described limitations of the published literature, the present study focused on two primary aims using PRSs for AUD that combine the effects of genetic variants across the genome into an aggregate score indexing genetic liability for AUD. First, we examined the relations between the AUD PRS and each AUD symptom to determine whether increases in the AUD PRS corresponded to endorsement of AUD symptoms of increasing severity as determined by previous IRT studies (Aim 1). As an overall measure of genetic risk for AUD, it was assumed that the PRS would be positively correlated with the relative severity of individual AUD criteria, such that more severe AUD criteria would be associated with higher mean scores of the AUD PRS. Second, we used MIMIC models to explore whether the AUD PRS predicted variation in any of the AUD symptoms after accounting for the covariance between symptoms as represented by a unidimensional latent AUD factor (Aim 2). Based on prior studies, we hypothesized that the PRS would significantly predict several individual symptoms (e.g., *withdrawal* & *craving*) after statistically adjusting for unidimensional AUD symptomatology, implying that the genetic risk for AUD cannot be fully captured by a single dimension.

## Methods

### Discovery Samples

#### Alcohol Use Disorder PRS

The AUD PRS was calculated using GWAS summary statistics for alcohol use disorder, derived from a previous meta-analysis. Information about sample characteristics, genotyping, quality control is illustrated in the study by Miller and Gizer [35]. Briefly, meta-analytic samples included a GWAS of alcohol dependence from the Psychiatric Genomics Consortium (PGC; *n_effective_* = 26,853) [36] and two AUD GWAS: FinnGen Consortium Release 6 (FinnGenR6; *n_effective_* = 40,997) [37] and Million Veteran Program (MVP; *n_effective_* = 152,332) [38]. A one-stage sample-size-weighted meta-analysis was conducted using METAL [39] for variants with a minor allele frequency (MAF) > 0.01 (9,533,157 variants). Only participants of European ancestry were included (*n_effective_* = 220,182). Note that *n_effective_* is calculated as 4/(1/N_cases_+1/N_ctrls_) and used by METAL as sample size weights to account for the unequal numbers of cases and controls across studies.

### Target Sample

#### University of California-San Francisco (UCSF)

A subset of participants from the University of California – San Francisco Family Alcoholism Study (UCSF) [40] were selected for this study. The UCSF Family Study recruited proband participants who met lifetime alcohol dependence criteria [41] and their available family members. Previous studies have found that the predictive ability of polygenic risk scores is decreased if there are differences in the ancestral backgrounds of participants in the target sample and discovery samples that result from differences in linkage disequilibrium (LD) patterns that are present across ancestry [42,43]. Therefore, participants of non-European ancestry were excluded from the UCSF target sample given that the discovery GWAS were conducted in European ancestry samples. Lastly, those who had never been exposed to alcohol were excluded, and only participants with both genetic and phenotypic data were included, yielding a total sample size of 1639 (1022 females and 617 males, mean age=49.69 [*SD*=12.92]). A total of 995 (60.86%) of the participants were formally diagnosed with DSM-IV Alcohol Dependence [44]. Institutional Review Board committees approved all data collection, and participants provided informed consent prior to participation.

#### Measures

The 11 AUD symptom criteria specified by DSM-5 were approximated using a set of items from a modified version of the Semi-Structured Assessment for the Genetics of Alcoholism (SSAGA) [45]. Craving was assessed by the item “In situations where you couldn’t drink, did you ever have such a strong desire for it that you couldn’t think of anything else” [22], thus allowing for the assessment of all 11 DSM-5 criteria (*tolerance, craving, larger/longer, cut/stop, role failure, interpersonal problems, time spent, reduced activities, withdrawal, hazardous use, and continued use [despite physical/psychological problems]*).

#### Whole genome sequencing

The sequencing and genotyping procedures for the UCSF Family Study have previously been detailed [46]. HiSeq2000 sequencers (Illumina, San Diego, CA) were used with most of the sample sequenced at a coverage depth between 2x and 6x. The LD-aware variant caller Thunder [47], which leverages the relatedness of the participants in the sample to improve genotype call accuracy, was used to make variant calls. The sample was also genotyped using Affymetrix microarrays to evaluate the quality of the genotype calls. Comparisons across platforms yielded a match rate of 98%. Principal component analysis (PCA) [48] was conducted using sequenced variants with a minor allele frequency (MAF) > 0.01 as implemented in the Genome-wide Complex Trait Analysis software (GCTA) [49].

#### Polygenic Risk Score (PRS)

The GWAS summary statistics from the AUD GWAS meta-analysis described above were used to construct the AUD PRS using PRS-CS [50], a Bayesian approach that models local LD patterns to produce posterior effect sizes of genetic variants that are conditioned on the non-independence of physically proximal variants. European samples from the 1000 Genomes project (phase 3, NCBI GRCh37) were used as a reference panel to model the LD between variants. Variants present across the discovery datasets, the LD reference panel, and the UCSF dataset were included. The PRS was then constructed by summing up weighted allele counts and standardizing them (mean = 0, *SD* = 1) using PLINK 2.0 [51].

### Data Analysis

To address Aim 1, mean values of the PRS of each AUD symptom were calculated by extracting PRS values for participants who endorsed the corresponding AUD symptom. For example, the mean PRS for *withdrawal* was calculated by averaging PRS values of participants who endorsed the *withdrawal* symptom. These mean values were then ordered from the highest to the lowest. This order was compared to the order of severity indices from a published meta-analysis of AUD IRT studies [52] by conducting a Spearman rank-order correlation analysis using the R *stats* package (version 4.3.1, R core team 2023).

To address Aim 2, a series of Multiple Indicators Multiple Causes (MIMIC) latent variable Item Response Theory (IRT) models were used to test the effects of the PRS on individual symptoms after accounting for their relation to a latent AUD variable. The MIMIC model is a special case of a structural equation model (SEM) comprised of two parts. One part is a measurement model where the relationship between a latent variable (i.e., AUD factor) and its indicators (i.e., individual AUD symptoms) is defined. The other part refers to a structural model where the relationship between a latent variable and observed background variables (i.e., PRS) are specified [53]. MIMIC models can include direct paths between observed background variables and latent variable indicators. These direct paths show differences in the tested indicator variables attributable to the observed background variables after statistically adjusting for the latent variable. In the present case, if the PRS (observed background variable) is significantly related to *withdrawal* (latent variable indicator), the degree of *withdrawal* is dependent on an individual’s PRS score even after the AUD factor score is covaried out (see Figure 1). In this way, we can test the measurement invariance of each AUD symptom across the PRS. Before constructing the MIMIC models, an exploratory factor analysis was conducted to check the dimensionality of AUD. As genetic relatedness within families could bias study results, observations were modeled as clustered within families to address the nonindependence among observations. Sex, mean-centered age, 10 ancestry principal components, and PRS were modeled as independent observed background variables predicting the AUD factor score in the MIMIC model. Then, the AUD PRS was regressed onto the individual symptom criteria. Because a model including paths from the PRS to all 11 indicators would not be identified, a set of 11 MIMIC models where the direct path from the PRS to a single AUD symptom was dropped in turn from the model were fit, and a *Bonferroni* corrected *p* < .005 was used to evaluate significance.

**Figure 1.**
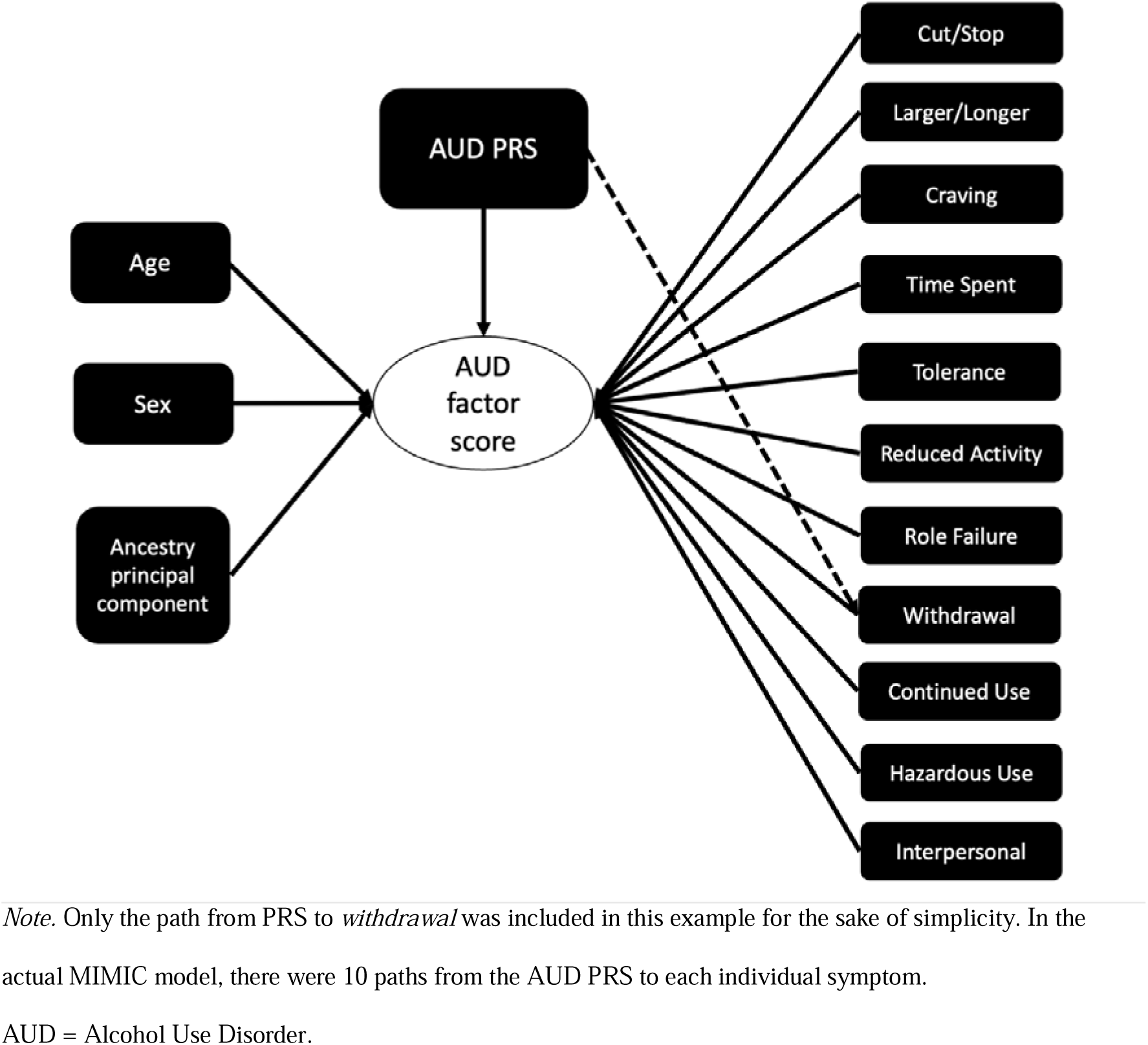
Example of the Multiple Indicator, Multiple Cause (MIMIC) model for withdrawal.

Analyses were conducted in Mplus version 8 [54]. Differences in model fit were evaluated using the chi-square difference test as well as the root mean squared error of approximation (RMSEA). For RMSEA, values < 0.08 were considered adequate. The Comparative Fit Index (CFI) and Tucker-Lewis Index (TLI) were also used, with values > 0.90 similarly indicating adequate model fit.

## Results

### Aim 1: Comparing AUD Symptom Severities at the Genetic and Phenotypic Levels

Mean PRSs among those who endorsed each AUD symptom ranged from 0.07-0.14 with *craving* showing the highest mean score and *cut/stop* the lowest. Table 1 displays the mean PRS among individuals endorsing each AUD symptom in descending order alongside corresponding IRT severity values (standardized θ). The rank order of mean PRSs was highly correlated with the rank order of median IRT severity indices (*r* = 0.78, *p* < 0.01) obtained from the cited meta-analyses [52].

**Table 1.** Order of PRS and IRT Severity Indices for Individual AUD Symptoms (from Most to Least Severe)

| Order | Severity indices |  |
| --- | --- | --- |
|  | PRS<br>Mean (% endorsed*) | IRT severity<br>Median |
| 1 | *Craving*<br>0.140 (37%) | *Reduced Activities*<br>0.610 |
| 2 | *Withdrawal*<br>0.139 (45%) | *Withdrawal*<br>0.571 |
| 3 | *Reduced Activities*<br>0.125 (41%) | *Craving*<br>0.295 |
| 4 | *Time Spent*<br>0.117 (41%) | *Role Failure*<br>0.161 |
| 5 | *Role Failure*<br>0.111 (44%) | *Time Spent*<br>0.100 |
| 6 | *Tolerance*<br>0.086 (60%) | *Interpersonal Problems*<br>0.099 |
| 7 | *Interpersonal Problems*<br>0.081(61%) | *Continued Use*<br>-0.106 |
| 8 | *Continued Use*<br>0.078 (62%) | *Cut/Stop*<br>-0.230 |
| 9 | *Larger/Longer*<br>0.076 (63%) | *Hazardous Use*<br>-0.464 |
| 10 | *Hazardous Use*<br>0.070 (59%) | *Tolerance*<br>-0.743 |
| 11 | *Cut/Stop*<br>0.069 (58%) | *Larger/Longer*<br>-0.953 |

*Note.* \*Endorsement rate of the corresponding symptom in UCSF sample.
Bold line indicates more than three rank difference.

### Aim2: Testing for Symptom-Specific PRS Effects

#### Exploratory Factor Analysis (EFA)

An EFA was conducted to evaluate one- and two-factor models. Both models showed good fit (one-factor model: RMSEA = .04, CFI= .99, TLI = .99; two-factor model: RMSEA = .03, CFI= 1, TLI = .99). The chi-square difference test was significant (χ^2^ = 85.87, df = 10, *p* < .001), suggesting preference for a two-factor model; however, examination of eigenvalues revealed that the second factor yielded an eigenvalue less than 1. Thus, the more parsimonious single-factor model was preferred (χ^2^ = 106.96, df = 44, *p* < .001), given the absence of consistent and interpretable associations with the second factor. This further supports the uni-dimensional treatment of AUD symptoms, and the further examination of measurement invariance of genetic risk for AUD in the subsequent MIMIC models.

#### MIMIC model

All 11 models iteratively dropping the path between the PRS and one AUD symptom in turn exhibited good fit (all chi-square *p*s < .001, RMSEAs < 0.08, CFIs > 0.90, TLIs > 0.90). Results of the MIMIC models indicated that the AUD PRS (see Figure 2), age, and sex significantly predicted the AUD factor score (all *p*s < .005), but none of the direct paths from the AUD PRS to AUD symptoms were significant (all *p*s > .118). Thus, there was no evidence suggesting unique effects of the PRS on individual AUD symptoms, independent of the latent AUD factor (See Table 2; see Supplementary Table 1 for results of other models).

**Figure 2.**
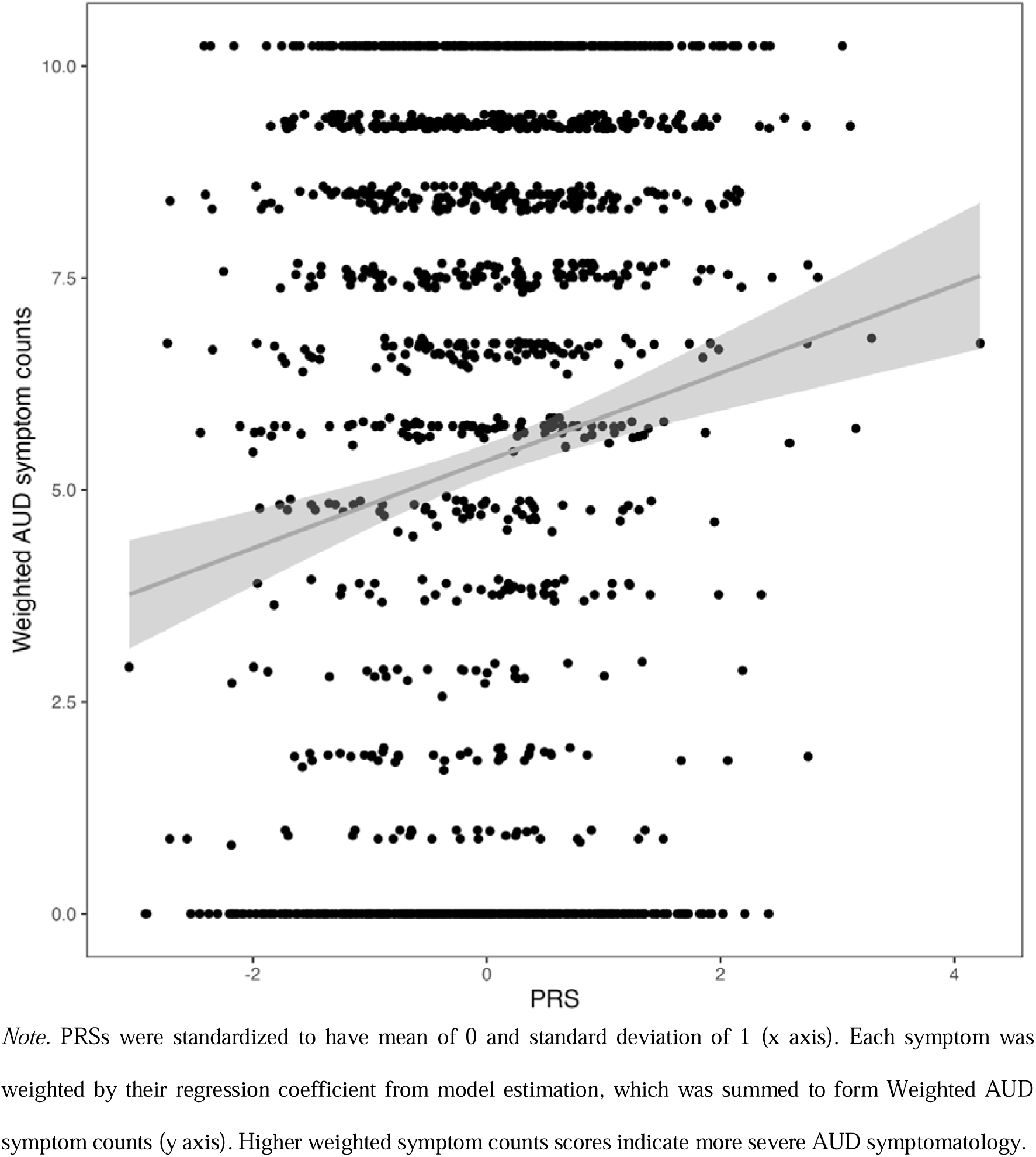
The Effect of AUD Polygenic Risk Scores (PRS) on the Weighted AUD Symptom Counts.

**Table 2.**
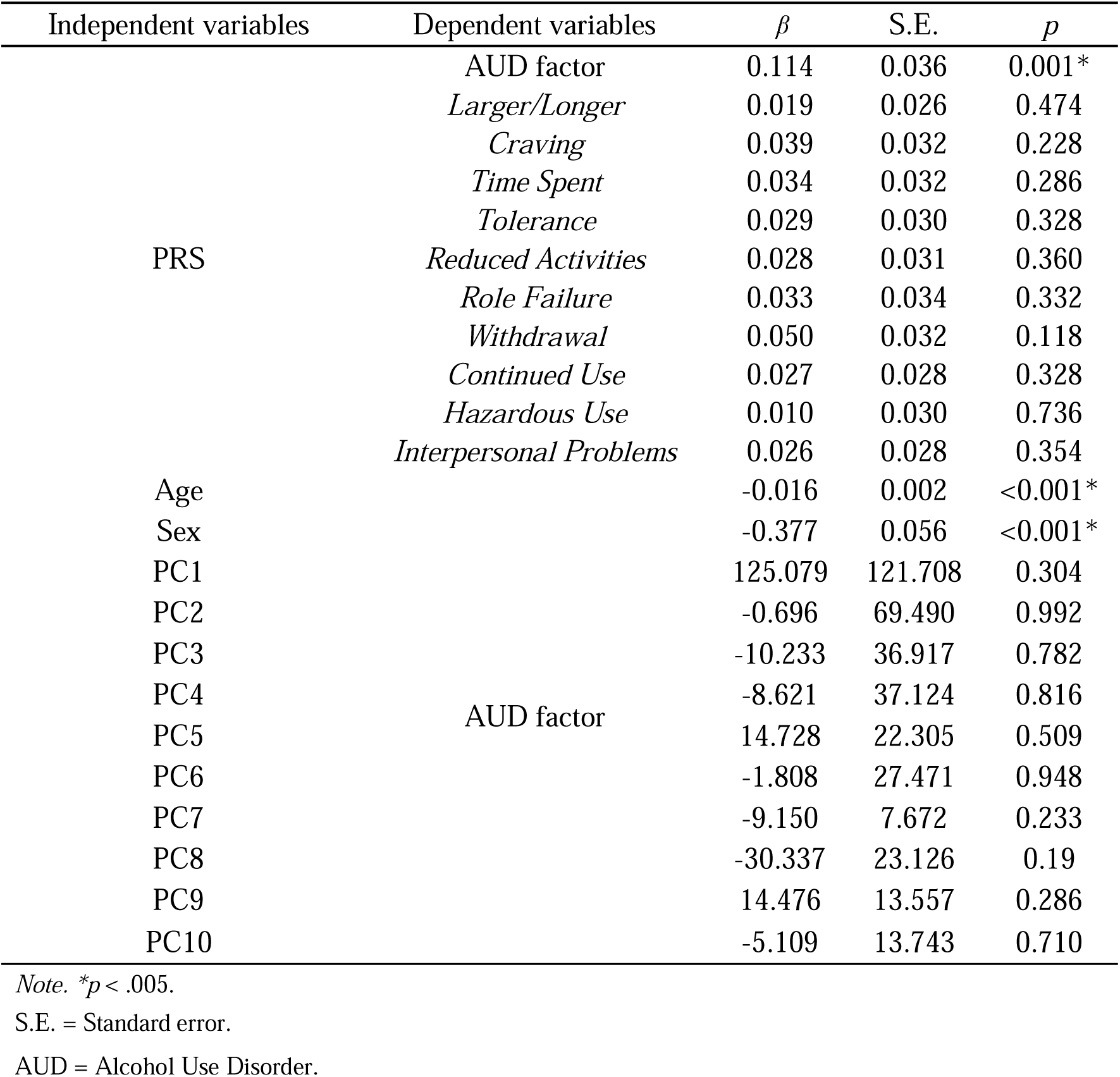
MIMIC Model Results of PRS and Covariates on AUD Factor Scores as well as PRS on each AUD Symptom (Model Without Cut/Stop Symptom)

| Independent variables | Dependent variables | $\beta$ | S.E. | $p$ |
| --- | --- | --- | --- | --- |
| PRS | AUD factor | 0.114 | 0.036 | 0.001* |
|  | *Larger/Longer* | 0.019 | 0.026 | 0.474 |
|  | *Craving* | 0.039 | 0.032 | 0.228 |
|  | *Time Spent* | 0.034 | 0.032 | 0.286 |
|  | *Tolerance* | 0.029 | 0.030 | 0.328 |
|  | *Reduced Activities* | 0.028 | 0.031 | 0.360 |
|  | *Role Failure* | 0.033 | 0.034 | 0.332 |
|  | *Withdrawal* | 0.050 | 0.032 | 0.118 |
|  | *Continued Use* | 0.027 | 0.028 | 0.328 |
|  | *Hazardous Use* | 0.010 | 0.030 | 0.736 |
|  | *Interpersonal Problems* | 0.026 | 0.028 | 0.354 |
| Age | AUD factor | -0.016 | 0.002 | <0.001* |
| Sex |  | -0.377 | 0.056 | <0.001* |
| PC1 |  | 125.079 | 121.708 | 0.304 |
| PC2 |  | -0.696 | 69.490 | 0.992 |
| PC3 |  | -10.233 | 36.917 | 0.782 |
| PC4 |  | -8.621 | 37.124 | 0.816 |
| PC5 |  | 14.728 | 22.305 | 0.509 |
| PC6 |  | -1.808 | 27.471 | 0.948 |
| PC7 |  | -9.150 | 7.672 | 0.233 |
| PC8 |  | -30.337 | 23.126 | 0.19 |
| PC9 |  | 14.476 | 13.557 | 0.286 |
| PC10 |  | -5.109 | 13.743 | 0.710 |

*Note.* \* $p < .005$ .
S.E. = Standard error.
AUD = Alcohol Use Disorder.

## Discussion

The primary aims of the present study focused on the use of polygenic scores to investigate potential genetic heterogeneity within the AUD diagnostic category in relation to severity of symptoms and the possibility of a multidimensional structure. First, we compared the genetic severity to the phenotypic severity of individual AUD symptoms and found a high degree of consistency. Second, we used MIMIC models to evaluate whether genetic influences underlying AUD were fully captured by a uni-dimensional latent factor or whether there were observed residual relations with individual symptoms suggesting symptom-specific genetic influences that, if correlated, could suggest a multi-dimensional structure. Findings supported the presence of a uni-dimensional genetic structure underlying AUD symptomatology in this sample. These findings have specific implications for understanding AUD assessment and diagnosis as well as future molecular genetics studies of AUD.

In terms of the findings from Aim 1, results demonstrating consistency between severities of individual AUD symptoms measured at the genotypic and phenotypic level add to the prior literature by demonstrating that the observed severity differences in phenotypic studies described in IRT studies have, at least in part, a biological basis. Similar to phenotypic studies [e.g., 33,52], we found that *withdrawal* and *craving* were associated with the highest level of genetic risk for AUD, symptoms related to social dysfunction, such as *role failure* and *interpersonal problems*, were associated with moderate genetic risk, and symptoms reflective of excessive consumption such as *larger/longer* were associated with the lowest level of genetic risk (see Table 1). Nonetheless, some discrepancies between level of genetic risk and phenotypic severity were noted. For example, *tolerance* was shown to be associated with a moderate level of genetic risk whereas phenotypic studies have consistently suggested that tolerance is one of the symptoms with the lowest severity [5,33,34]. Nonetheless, the high degree of similarity across genotypic and phenotypic measures of severity is encouraging in its support for ongoing GWAS of AUD by suggesting that the genetic variants being identified in these studies are in fact reflective of the operationalization of the disorder as described in the DSM. Put more simply, phenotype definitions matter. Notably, this may be viewed as both a positive and negative in relation to AUD as the DSM-5 AUD diagnosis has both supporters and detractors [e.g., 6,31].

Of direct importance to the field of alcohol research and conceptualizations of AUD, the findings from the present study suggest that differences in AUD severity attributed to individual AUD symptoms in IRT studies are at least partially a reflection of varying levels of genetic risk. A direct implication of this finding is that the current DSM practice of treating all AUD symptoms as equal indicators of disease may not be ideal, a position that has been argued for in the literature. For example, Lane and Sher [28] used data from NESARC-II to compare different symptom profiles of individuals reporting symptoms of AUD and found that certain symptom combinations such as *reduced activities* and *role failure* were indicative of a more severe form of AUD (e.g., drinking behavior & physical and mental health) relative to other symptom combinations, thus concluding that symptom configurations, rather than symptom counts, may be more valid indicators of AUD severity, a finding that was replicated by [30] using data from the Collaborative Study on the Genetics of Alcoholism (COGA). The present study suggests a similar conclusion in that endorsements of more severe AUD symptoms are reflective of increasing levels of genetic risk for AUD. Together, these results suggest that there may be meaningful differences across AUD symptoms in terms of their severity that have relevance to prevention and intervention efforts by identifying more at-risk individuals or more severe AUD cases in greater need of services than is reflected in a symptom count. Thus, weighting all AUD symptoms as equal when considering an AUD diagnosis may come at the cost of losing important diagnostic information relevant to prognosis.

The second major implication of the present study comes from the largely null results of the MIMIC models in identifying residual genetic influences on AUD symptoms after accounting for the general latent factor. The presence of such influences would have suggested a multidimensional genetic architecture underlying AUD. Our null results thus support the conclusion that AUD genetic liability is unidimensional, which was contrary to our expectation, but importantly, do not preclude a multidimensional structure, a position supported by prior animal and human studies [e.g., 7,14]. Further, while other studies have produced evidence suggesting that AUD might be comprised of a single genetic factor, those studies have typically reported symptom-specific genetic influences as well [e.g., 18,19], a finding that has been supported across multiple, distinct methodologies [e.g., 22,24].

One potential, and likely, explanation for the present study’s support of a unidimensional genetic structure of AUD is the nature of the GWAS summary statistics used to create the AUD PRS. These summary statistics were created from GWAS of AUD diagnostic status, which assumes a unidimensional phenotypic structure. This could have biased results towards a single-factor genetic structure of AUD when tested using AUD PRS as was done in this study. The high level of correspondence between the genotypic and phenotypic measures of individual symptom severities found in the present study supports this conclusion in providing a demonstration of how phenotype definitions can be reflected back in GWAS findings. Thus, given the breadth of studies that have argued for a multi-dimensional conceptualization of AUD [e.g., 5-7], the present study suggests that data from item-level GWAS will likely be required to rigorously test multidimensional models of AUD at the genetic level. Though not a study of AUD symptoms, one such study conducted an item-level GWAS of the Alcohol Use Disorder Identification Test (AUDIT) to demonstrate a two-dimensional factor-structure of that measure [55]. The present study strongly suggests that item-level GWAS of AUD symptoms will be required to more thoroughly investigate the genetic factor structure of AUD, a conclusion that holds for psychiatric disorders more broadly and one that requires more attention in the field.

There are some limitations of this study that should be considered. First, the data in our study were restricted to participants of European ancestry, which limits the generalizability of the results. Because of variation in linkage disequilibrium patterns among individuals that differ in genetic ancestry, early GWAS tended to focus on a single ancestry, typically European ancestry [56]. Although trans-ancestry GWAS have now been conducted for AUD, the low number of non-European ancestry participants in the UCSF study sample did not allow us to include these participants in the present study. Second, we assumed uni-dimensionality for the AUD latent variable. Although the data supported a one-factor solution, there was some limited evidence supporting a two-factor solution. Nonetheless, the degree of support for a one-factor vs. two-factor solution was similar to the many previously published IRT studies of AUD symptoms [e.g., 34,57].

In summary, the present study is the first attempt to utilize the PRS approach to examine the relations between AUD symptom severities and AUD genetic risk. Our findings demonstrate that the severities of AUD symptoms in relation to an underlying AUD trait are partly a reflection of increasing genetic risk and that there is minimal evidence suggesting the AUD PRS is capturing symptom-specific genetic influences operating outside of this underlying AUD trait. These findings illustrate how GWAS of AUD have contributed to the field, but also demonstrate that if we take the conclusions from phenotypic studies suggesting that AUD symptomatology may be multidimensional seriously, future studies will need to conduct item-level GWAS of AUD symptoms to test these alternative structures at the genetic level.

## Data Availability

Genotype data for the UCSF Family Study can be obtained through the Database for Genotypes and Phenotypes (dbGaP; Study Accession: phs001458.p1.v1). Phenotype data are available from the corresponding author upon request.PGC alcohol dependence GWAS summary statistics were obtained from the PGC website (https://www.med.unc.edu/pgc/). Million Veteran Program GWAS summary statistics were obtained through the Database for Genotypes and Phenotypes (dbGaP; Study Accession: phs001672). FinnGenR6 ICD-based AUD GWAS data were obtained from https://www.finngen.fi/en/access_results.

https://www.finngen.fi/en/access_results

## Funding

The UCSF Family Alcoholism Study was funded in part by the National Institute on Drug Abuse: R01DA030976 (principal investigator: Kirk C. Wilhelmsen), and the State of California for medical research on alcohol and substance abuse through the University of California at San Francisco and Ernest Gallo Clinic and Research Center to Kirk C. Wilhelmsen. National Institute of Alcohol Abuse and Alcoholism: F31AA027957 (principal investigator: Alex P. Miller).

## Acknowledgments

This study made use of summary statistics data from a number of sources. First, this study used GWAS summary statistics data from the fifth release of the FinnGen study. We thank the participants and investigators of the FinnGen study. Second, this research used summary data from the Psychiatric Genomics Consortium Substance Use Disorders (PGC-SUD) working group. PGC–SUD is supported by funds from NIDA and NIMH to MH109532 and, previously, had analyst support from NIAAA to U01AA008401 (COGA). PGC–SUD gratefully acknowledges its contributing studies and the participants in those studies, without whom this effort would not be possible. Third, this research used data from the Million Veteran Program and was supported by funding from the Department of Veterans Affairs Office of Research and Development, Million Veteran Program Grant nos. I01BX003341 and I01CX001849; and the VA Cooperative Studies Program study, no. 575B. This publication does not represent the views of the Department of Veterans Affairs or the United States Government.

## Notes

We have no conflicts of interests to disclose.

### Competing Interest Statement

The authors have declared no competing interest.

### Author Declarations

The University of California San Francisco and the University of North Carolina at Chapel Hill institutional review boards provided ethical approval for all data collection procedures.

