## Supplementary Table for "Genetic Risk for Alcohol Use Disorder in Relation to Individual Symptom Criteria: Do Polygenic Indices Provide Unique Information for Understanding Severity and Heterogeneity?"

| Supplementary Table 1 | | | | | | | | | | | | |
| --- | --- | --- | --- | --- | --- | --- | --- | --- | --- | --- | --- | --- |
| Full results of MIMIC models for the AUD PRS predicting the latent AUD factor and each AUD symptom. | | | | | | | | | | | | |
|  |  | MIMIC Model without ________ symptom | | | | | | | | | | |
| Dependent variables | Statistics | Cut/Stop | Lager/longer | Craving | Time spent | Tolerance | Activities given up | Role failure | Withdrawal | Continued use | Hazardous use | Interpersonal problems |
| AUD factor | *β* | 0.114 | 0.134 | 0.156 | 0.156 | 0.145 | 0.145 | 0.153 | 0.165 | 0.142 | 0.126 | 0.140 |
|  | S.E. | 0.036 | 0.034 | 0.037 | 0.043 | 0.036 | 0.037 | 0.038 | 0.034 | 0.036 | 0.039 | 0.036 |
|  | *p* | 0.001* | <0.001* | <0.001* | <0.001* | <0.001* | <0.001* | <0.001* | <0.001* | <0.001* | 0.001* | <0.001* |
| Cut/Stop | *β* | - | -0.018 | -0.038 | -0.039 | -0.028 | -0.029 | -0.036 | -0.047 | -0.025 | -0.011 | -0.024 |
|  | S.E. | - | 0.025 | 0.033 | 0.037 | 0.029 | 0.031 | 0.037 | 0.030 | 0.026 | 0.032 | 0.026 |
|  | *p* | - | 0.475 | 0.245 | 0.298 | 0.332 | 0.365 | 0.341 | 0.122 | 0.332 | 0.737 | 0.358 |
| Lager/ longer | *β* | 0.019 | - | -0.022 | -0.023 | -0.011 | -0.012 | -0.019 | -0.031 | -0.008 | 0.008 | -0.007 |
|  | S.E. | 0.026 | - | 0.034 | 0.038 | 0.024 | 0.033 | 0.036 | 0.030 | 0.020 | 0.032 | 0.021 |
|  | *p* | 0.474 | - | 0.527 | 0.550 | 0.644 | 0.728 | 0.599 | 0.303 | 0.698 | 0.813 | 0.746 |
| Craving | *β* | 0.039 | 0.021 | - | -0.001 | 0.010 | 0.010 | 0.003 | -0.009 | 0.013 | 0.028 | 0.015 |
|  | S.E. | 0.032 | 0.032 | - | 0.039 | 0.034 | 0.034 | 0.039 | 0.032 | 0.033 | 0.039 | 0.033 |
|  | *p* | 0.228 | 0.519 | - | 0.983 | 0.756 | 0.771 | 0.947 | 0.782 | 0.684 | 0.472 | 0.657 |
| Time spent | *β* | 0.034 | 0.019 | 0.001 | - | 0.010 | 0.009 | 0.003 | -0.007 | 0.012 | 0.025 | 0.013 |
|  | S.E. | 0.032 | 0.031 | 0.033 | - | 0.032 | 0.028 | 0.033 | 0.032 | 0.031 | 0.035 | 0.031 |
|  | *p* | 0.286 | 0.546 | 0.984 | - | 0.766 | 0.749 | 0.930 | 0.826 | 0.699 | 0.480 | 0.673 |
| Tolerance | *β* | 0.029 | 0.011 | -0.011 | -0.012 | - | -0.001 | -0.008 | -0.020 | 0.003 | 0.018 | 0.004 |
|  | S.E. | 0.030 | 0.023 | 0.035 | 0.039 | - | 0.033 | 0.038 | 0.030 | 0.023 | 0.031 | 0.024 |
|  | *p* | 0.328 | 0.644 | 0.758 | 0.767 | - | 0.984 | 0.833 | 0.508 | 0.899 | 0.560 | 0.860 |
| Activities given up | *β* | 0.028 | 0.011 | -0.009 | -0.010 | 0.001 | - | -0.007 | -0.018 | 0.003 | 0.018 | 0.005 |
|  | S.E. | 0.031 | 0.030 | 0.033 | 0.032 | 0.031 | - | 0.029 | 0.030 | 0.030 | 0.036 | 0.030 |
|  | *p* | 0.360 | 0.727 | 0.773 | 0.751 | 0.984 | - | 0.812 | 0.555 | 0.911 | 0.623 | 0.879 |
| Role Failure | *β* | 0.033 | 0.016 | -0.002 | -0.003 | 0.007 | 0.006 | - | -0.010 | 0.010 | 0.023 | 0.011 |
|  | S.E. | 0.034 | 0.031 | 0.035 | 0.034 | 0.033 | 0.027 | - | 0.032 | 0.031 | 0.036 | 0.031 |
|  | *p* | 0.332 | 0.596 | 0.947 | 0.930 | 0.832 | 0.811 | - | 0.742 | 0.755 | 0.524 | 0.729 |
| Withdrawal | *β* | 0.050 | 0.031 | 0.009 | 0.008 | 0.020 | 0.019 | 0.012 | - | 0.023 | 0.038 | 0.024 |
|  | S.E. | 0.032 | 0.030 | 0.033 | 0.038 | 0.030 | 0.033 | 0.036 | - | 0.030 | 0.036 | 0.030 |
|  | *p* | 0.118 | 0.298 | 0.779 | 0.825 | 0.503 | 0.552 | 0.741 | - | 0.445 | 0.285 | 0.415 |
| Continued use | *β* | 0.027 | 0.008 | -0.014 | -0.015 | -0.003 | -0.004 | -0.011 | -0.023 | - | 0.016 | 0.001 |
|  | S.E. | 0.028 | 0.021 | 0.035 | 0.039 | 0.023 | 0.033 | 0.036 | 0.031 | - | 0.032 | 0.007 |
|  | *p* | 0.328 | 0.697 | 0.689 | 0.702 | 0.899 | 0.912 | 0.757 | 0.449 | - | 0.624 | 0.852 |
| Hazardous use | *β* | 0.010 | -0.007 | -0.026 | -0.027 | -0.017 | -0.017 | -0.024 | -0.035 | -0.014 | - | -0.013 |
|  | S.E. | 0.030 | 0.029 | 0.037 | 0.039 | 0.028 | 0.035 | 0.038 | 0.033 | 0.029 | - | 0.029 |
|  | *p* | 0.736 | 0.813 | 0.480 | 0.490 | 0.560 | 0.627 | 0.533 | 0.298 | 0.627 | - | 0.658 |
| Interpersonal problem | *β* | 0.026 | 0.007 | -0.015 | -0.016 | -0.004 | -0.005 | -0.012 | -0.025 | -0.001 | 0.014 | - |
|  | S.E. | 0.028 | 0.021 | 0.035 | 0.038 | 0.024 | 0.032 | 0.036 | 0.031 | 0.007 | 0.032 | - |
|  | *p* | 0.354 | 0.745 | 0.662 | 0.675 | 0.861 | 0.879 | 0.732 | 0.420 | 0.852 | 0.656 | - |
| **p* < .005 | | | |  |  |  |  |  |  |  |  |  |
| S.E. = Standard error. | | | |  |  |  |  |  |  |  |  |  |
| AUD = Alcohol Use Disorder | | | |  |  |  |  |  |  |  |  |  |
